# Paraoxonase 1 status is a major Janus-faced component of mild and moderate acute ischemic stroke and consequent disabilities

**DOI:** 10.1101/2022.08.12.22278728

**Authors:** Francis F. Brinholi, Ana Paula Michelin, Andressa K. Matsumoto, Laura de O. Semeão, Abbas F. Almulla, Thitiporn Supasitthumrong, Chavit Tunvirachaisakul, Decio S. Barbosa, Michael Maes

## Abstract

**Aims:** This study aims to examine the associations between paraoxonase 1 (PON)1 status and acute ischemic stroke (AIS) and consequent disabilities.

**Methods:** This study recruited 122 patients with AIS and 40 healthy controls and assessed the Q192R gene variants, arylesterase (AREase) and chloromethyl phenylacetate (CMPAase) activities, and high-density lipoprotein cholesterol (HDL) in baseline conditions. AREase and CMPAase were measured 3 months later. The National Institutes of Health Stroke Scale (NIHSS) and the modified Rankin score (mRS) were assessed at baseline and 3 and 6 months later.

**Results:** Reduced CMPAase and increased AREase activities are significantly associated with AIS and mRS and NIHSS scores (baseline and 3 and 6 months later). The best predictor of AIS/disabilities was a decrease in the z-unit-based composite zCMPAase-zAREase score. Serum high density lipoprotein cholsterol (HDL) was significantly correlated with CMPAase, but not AREase, activity and a lowered zCMPAase+zHDL score was the second best predictor of AIS/disabilities. Regression analysis showed that 34.7% of the variance in baseline NIHSS was explained by zCMPAase-zAREase and zCMPAase+zHDL composites, HDL, and hypertension. Neural network analysis showed that stroke was differentiated from controls with an area under the ROC curve of 0.975 using both new composite scores, PON1 status, hypertension, dyslipidemia, previous stroke as body mass index. The PON1 Q192R genotype has many significant direct and mediated effects on AIS/disabilities, however, its overall effect was not significant.

**Discussion:** PON1 status and the CMPAase-HDL complex play key roles in AIS and its disabilities at baseline and 3 and 6 months later.

## Introduction

Stroke is the second leading cause of death and disability, with about 13 million new cases per year (GBD, 2019; Chen et al., 2020; World Stroke Organization, 2021). An estimated one in four persons may suffer a stroke in their lifetime, and there are more than 100 million stroke survivors worldwide (Campbell et al., 2019; World Stroke Organization, 2021). The expanding and aging population, according to projections, will lead this number to increase considerably over the next several decades (Stinear, Lang Zeiler, Byblow, 2020). Acute ischemic stroke (AIS), which accounts for 87% of all strokes worldwide, is described as the sudden loss of blood flow to an area of the brain resulting in the loss of neurologic function, caused by thrombosis or embolism that occludes a cerebral vessel serving a specific area of the brain (Wajngarten, Silva, 2019; Phipps, Cronin; 2020). Using the National Institutes of Health Stroke Scale (NIHSS) and the modified Rankin score (mRS) (Brott et al., 1989; Bonita, Beaglehole, 1988), the baseline AIS severity and AIS-induced impairments can be graded, respectively. Both scales are useful for predicting the functional result of AIS, either in the short-term (3 months) or long-term (6 months) (Adams et al., 1999; Johnston et al., 2002; Weimar et al., 2004).

AIS risk factors include unmodifiable factors such as age and gender, and modifiable factors such as diabetes mellitus, high systolic blood pressure, high body mass index, heart disease, high levels of low-density lipoprotein (LDL)-cholesterol and low levels of high density lipoprotein (HDL)-cholesterol, atrial fibrillation, transient ischemic attack (TIA), smoking, sedentary lifestyle and nutritional factors (Gorelick, 2019; Maes et al., 2021). AIS is accompanied by increased production of reactive oxygen and nitrogen species (RONS) and activated immune-inflammatory pathways that cause neuronal damage (Orellana-Urza et al., 2020; Maes et al., 2021). As a result, a portion of the brain tissue (core) sustains irreversible neuronal loss due to necrotic cell death, whereas the surrounding tissues contain recoverable and metabolically active cells (penumbra) in which cell death happens more slowly (Khoshnam et al., 2017). Additionally, AIS and AIS outcome are accompanied with peripheral indications of oxidative and nitrosative stress (O&NS), lowered antioxidant defenses and activated immune-inflammatory pathways, including C-reactive protein (CRP) (Lehmann et al., 2022; Alfieri et al., 2020).

Paraoxonases (PON) are an enzyme family comprising of PON1, PON2, and PON3 that hydrolyze organophosphate chemicals (Park et al., 2015; Kulka, 2016; Xu et al., 2021). PON1 is a plasmatic enzyme with paraoxonase and arylesterase (AREase) activity, as well as antioxidative and anti-inflammatory effects (Tisato et al., 2019; Bliźniewska-Kowalska et al., 2022). PON1 is an HDL-associated protein that shields HDL and LDL cholesterol from oxidation and can hydrolyze oxidized LDL-cholesterol, which may have atheroprotective properties (Varela et al., 2020). In addition, PON1 inhibits macrophage oxidative capacity, RONS generation, and cholesterol synthesis, hence exerting atheroprotective actions (Tajbakhsh et al., 2017). PON1 activity is sensitive to increased oxidative stress and inflammatory responses which may both decrease PON1 activities (Maes et al., 2022). Low PON1 activity has repeatedly been associated with an increased risk of atherosclerosis, which contributes to heart attack, stroke, and vascular problems (Zhang et al., 2013; Menini, Gugliucci, 2014; Park et al., 2015). Furthermore, compared to AIS patients with a better outcome, AIS patients with poor outcomes have decreased serum PON1 activity (Xu et al. 2021). After a vascular event, a decreased PON1 activity may be noticed, and this decrease may persist for an extended length of time (Tajbakhsh et al., 2017).

Two major polymorphisms in the PON1 coding area result in a glutamine (Q) arginine (R) substitution at position 192 (Q192R) in the PON1 gene (Yildiz et al., 2017; Gupta et al., 2022). This Q192R polymorphism may increase illness development risk and disease severity (Shunmoogam, Naidoo, Chilton, 2018) via altering the catalytic efficiency of PON1 (Tajbakhsh et al., 2017). The Q192 isoform hydrolyzes paraoxon more quickly, metabolizes oxidized LDL more efficiently, and inhibits LDL oxidation more effectively than the 192R isoform (Costa et al., 2005; Tajbakhsh et al., 2017). There is some evidence that the 192 R allele increases the risk of stroke in hypertensive individuals (Odds ratio=1.25, 95% confidence intervals: 1.07, 1.46) (Benerjee, 2010 et al., Dahabreh et al., 2010), although some studies have yielded negative results (Gupta et al., 2022).

However, it is difficult to interpret the effects of PON1 gene variants without considering the activities of different PON1 catalytic sites, namely AREase versus chloromethyl phenylacetate (CMPAase) activities, due to effects of “indirect-only mediation” (Maes et al., 2022). For example, in epilepsy, CMPAase, but not AREase, activity mediates the effects of the PON1 gene on the epilepsy phenome even without direct effects of the gene variants may be observed. Moreover, it should be emphasized that collinearity issues make the statistical analysis of PON1 status data challenging (Maes et al., 2022).

Hence, this study delineates the associations between stroke and disabilities (as assessed with mRS and NIHSS scores) and PON1 status as determined by the Q192R PON1 gene variants, CMPAase, AREase activity, the residualized CMPAase and AREase activities (to examine of the effects of the catalytic sites independently from the genetic variants), the composite of zCMPAase+zHDL cholesterol (to examine the antioxidant capacity of the PON1-HDL complex).

## Subjects and Methods

### Participants

This study recruited 122 patients with acute cerebral infarction hospitalized at the Stroke Unit of King Chulalongkorn Memorial Hospital between October 2019 and September 2020. Eligible patients had acute ischemic stroke and focal neurological signs or symptoms of vascular origin that persisted longer than 24 hours, as verified by clinical examination by a senior neurologist and confirmed by brain CT scan measurements. Part of the patients were again screened 3 and 6 months later. Normal controls (n=40) were obtained by word of mouth from the same catchment region (Bangkok province, Thailand). Exclusion criteria for patients were as follows: a) patients who had a hemorrhagic stroke or transient ischemic attack; b) patients who were unconscious, aphasic, or had severe cognitive impairments and were unable to cooperate with the verbal clinical interview; and c) patients who failed to perform MRI scans. The exclusion criteria for patients and controls were a) major medical disorders such as (auto)immune illness including rheumatoid arthritis, psoriasis, type 1 diabetes mellitus, systemic lupus erythematosus, inflammatory bowel disease, COPD, renal or liver failure, cancer, b) neuroinflammatory and neurodegenerative disorders including multiple sclerosis, Alzheimer’s and Parkinson’s; c) infectious disease including HIV and hepatitis B infection; d) premorbid axis-1 psychiatric DSM-5 diagnoses such as a major depressive episode, bipolar disorder, schizophrenia, post-traumatic stress disorder, post-traumatic stress disorder, and e) substance-abuse and psycho-organic disorders and use of antidepressants or other psychoactive drugs. Any axis-1 mental illness and a positive family history of depression and mood disorders in first-degree relatives were exclusion criteria for controls.

The study was approved by the Institutional Review Board of Chulalongkorn University’s Faculty of Medicine in Bangkok, Thailand (IRB no. 62/073), which is in accordance with the International Guideline for Human Research Protection, as required by the Declaration of Helsinki, the Belmont Report, the CIOMS Guidelines, and the International Conference on Harmonization in Good Clinical Practice (ICH-GCP). All participants, as well as the guardians of the patients, gave written informed consent prior to inclusion in the study.

### Clinical measurements

Sociodemographic data such as age, gender, years of education, marital status, employment, and vascular risk factors such as dyslipidaemia, type 2 diabetes mellitus (T2DM), hypertension, prior stroke, ischemic heart disease, and atrial fibrillation were collected by semi-structured interviews. All participants had the National Institutes of Health Stroke Score (NIHSS) and Modified Rankin Scale (mRS) scores evaluated, in patients within 8 hours of admission (baseline values). Both scales were reassessed after 3- and 6-months follow-up in part of the patients. The mRS is commonly used in contemporary clinical practice and indicates the degree of disability, global impairment, or dependence after a stroke. (Banks and Marotta, 2007). The mRS is scored as no symptoms at all (0 score), some symptoms but no disability (1), and slight (2), moderate (3), moderately severe (4) and severe disability (5). The mRS scores categorize the stroke outcome as good, namely mRS <3, and poor, namely mRS ≥3 (Park et al., 2015). The NIHSS quantifies stroke severity by assessing key domains such as levels of consciousness, visual fields, extraocular movements, facial palsy, arm and leg motor drift, limb ataxia, sensation, language, dysarthria and inattention (The National Institute of Neurological Disorders and Stroke rt-PA Stroke Study Group, 1995). A high NIHSS score indicates more impairment in physical functioning after a stroke (Alajbegovic et al., 2014) with an NIHSS score 1-4 indicating minor stroke and 5-10 moderate stroke. In the present study we have constructed an overall index of severity of stroke as z transformation of the mRS score (z mRS) + z NIHSS. In part of the patients, both the mRS and NIHSS examinations were repeated three and six months later. The TOAST criteria were used to establish the ischemic stroke subtype, which includes lacunar infarction (LAC), cardioembolic infarction (CEI), large artery atherosclerosis (LAAS), stroke of other determined aetiology (ODE) and stroke of unknown aetiology (UDE) (Adams et al., 1993).

## Assays

The present study defined PON1 status as the PON1 Q192R genotypes, and CMPAase and AREase activities (Moreira al., 2019a; Maes et al., 2022; Matsumoto et al., 2021). After an overnight fast, blood samples were obtained at 8:00 a.m., and serum was aliquoted and kept at -80 °C until thawing to determine PON1 status. Total PON1 activity was evaluated by phenylacetate hydrolysis formation (Moreira et al., 2019a). The rate of phenylacetate hydrolysis was assessed using a Perkin Elmer® EnSpire model microplate reader (Waltham, MA, USA) at a wavelength of 270 nm for a period of 4 minutes (16 readings with 15 seconds between each reading) at a constant temperature of 25°. Based on the phenyl acetate molar extinction coefficient of 1.31 mMol/Lcm^-1^, the activity was expressed in units per milliliter (U/mL). We studied the rate of phenylacetate hydrolysis at low salt concentration assaying arylesterase (AREase) as well as chloromethyl phenylacetate (CMPA, Sigma, USA)-ase activity (Moreira et al., 2019a; Maes et al., 2022; Matsumoto et al., 2021). We also employed CMPA and phenyl acetate to stratify the functional genotypes of the PON1Q192R polymorphism (PON1 192Q/Q, PON1 192Q/R, PON1 192R/R) (PA, Sigma, USA). PON1 polymorphism, which imparts variability in hydrolysis capability, allows to the stratification of genotypes based on phenotypic enzyme activities. Isoform R has a high hydrolysis activity on CMPA, while the Q alloenzyme possesses a reduced hydrolysis activity on CMPA. The phenylacetate reaction is conducted with high salt concentrations, which partly limits the activity of the R allozyme, allowing for a clearer separation between the three functional genotypes.

All of the controls and all of the patients at the start of the study had their CMPAase and AREase activity measured. Some of the patients had their activity measured again 3 months later. Moreover, we also computed the CMPAase/AREase activity ratio as zCMPAase–zAREase, which reflects the difference in activity between both catalytic sites of the PON1 enzyme. Finally, we also computed the zCMPAase+zHDL index, which reflects the total antioxidant capacity of the PON1+HDL complex. In order to compute the enzyme activity independent from the PON1 genotype we computed residualized CPMAase and AREase act ivities which have the effects of the genotype partialled out using regression analysis. High sensitivity C-Reactive Protein (hsCRP) was assayed employing a high sensitivity CRP Vario assay (Abbott Laboratories, Abbott Park, Illinois) on Architect cSystems. HDL cholesterol was assayed using an accelerator selective detergent on the Architect C8000 (Abbott Laboratories, Abbott Park, Illinois, USA). The intra-assay coefficient variation for both hsCRP and HDL was < 4.0%.

### Statistics

Contingency tables were used to evaluate the associations between categorical variables, and the Chi-square test or Fisher exact (Fisher-Freeman-Halton Exact) Test’ test was used where suitable. We used analysis of variance to investigate the differences in continuous variables across groups (ANOVA). Pearson’s product moment correlation coefficients were utilized to measure variable intercorrelations. To normalize the distribution of variables or to account for heterogeneity of variance across research groups (as assessed with the Levene test) continuous data were converted using logarithmic (Log) or square root transformations (e.g. CRP and NLR in Log transformation). Multiple regression analysis including with forward stepwise method were utilized to determine the most relevant biomarkers that predict rating scale scores while allowing for the effects of demographic variables. R^2^ variance, multivariate normality (Cook’s distance and leverage), homoscedasticity (as evaluated by the White and modified Breusch-Pagan tests), and (multi)collinearity as assessed with variance inflation factor (cut off >4), tolerance (<0.25) and the condition index and variance proportions in the collinearity diagnostics table were all explored. Where needed we group predictors for example by making composite scores (see the ratios as explained above) to solve collinearity problems or to reduce the number of features. We also used a stepwise automated technique with 0.05 p- to-entry and 0.06 p-to-remove. All regression analysis results were bootstrapped (5.000 samples), and the latter are shown if the findings do not agree. The statistical technique utilized to examine repeated measurements (from baseline to month 3, from the latter to month 6 and from baseline to month 3 and 6) was generalized estimating equations (GEE) with the NIHSS or mRS values from month 3 to month 6 as dependent variable and the changes in biomarkers from baseline to month 3 as explanatory variables while allowing for the effects of possible confounding variables (e.g. age, sex, BMI, comorbidities). We used multilayer perceptron neural network models to investigate the relationships between AIS and controls (output variables) and PON1 status data, classical risk factors, BMI, age, and sex. We used an automated feedforward architecture model with one to two hidden layers and trained it using maximal 8 nodes and 250 epochs, and minibatch training. The halting condition was one consecutive step with no further reduction in the error term. The subjects were separated into three groups: a) a training sample to estimate model parameters (46.67 percent of all cases); b) a testing sample to avoid overtraining (20% of all cases); and c) a holdout set to assess predictive validity (33.33 percent of all patients). The error, relative error, percentage of wrong classifications, area under receiver operating characteristic curve (AUC ROC), and (relative) relevance of the input variables were calculated. The confusion matrix table included the accuracy as well as sensitivity and specificity of the model. Automatic stepwise binary logistic regression analysis was performed with stroke as the dependent variable (reference group is controls) and using PON1 status, HDL, hsCRP, BMI, age, and sex as explanatory factors. The findings are reported as an adjusted odds ratio (OR) with a 95% confidence interval (CI), with Wald (W) statistics and the Nagelkerke pseudo R^2^ as effect size. All tests are two-tailed, with a significance level of p=0.05. All statistical analyses are carried out with the help of IBM SPSS Windows version 25, 2017.

Partial least squares (PLS) path analysis was used to investigate the causal relationships between the PON1 gene, hypertension, inflammation (CRP), the PON1 activities CMPAase and AREase activity, HDL cholesterol, and the phenome of stroke. The stroke phenome was entered as a latent vector, whereas all other input variables were entered as single indicators. PLS path analysis was performed only when the inner and outer models satisfied the following predetermined quality criteria: a) the output latent vector demonstrates accurate construct and convergence validity as indicated by average variance explained (AVE) > 0.5, Cronbach’s alpha > 0.7, composite reliability > 0.8, and rho A > 0.8, b) all outer loadings are > 0.6 at p 0.001, c) the construct’s cross-validated redundancy is good as tested using Blindfolding, c) the model’s prediction performance is adequate using PLSPredict, d) the overall model fit namely the standardized root square residual (SRMR) value is accurate with a value <0.08, and e) Confirmatory Tetrad Analysis (CTA) showed that the model is not misspecified as a reflective model. If all of the above-mentioned model quality data meet the predetermined criteria, we conduct a comprehensive PLS path analysis with 5,000 bootstrap samples, produce the path coefficients (with exact p-values), and additionally compute the specific and total indirect (that is, mediated) effects as well as the total effects.

## Results

### Demographic and clinical data

**Table 1** shows the socio-demographic and clinical data of the AIS patients and controls. We have divided the patient group into two different subgroups based on the z NIHSS + z mRS index using a threshold value of 0.82. One subgroup comprising 37 patients showed moderately increased NIHSS and mRS scores and in therefore dubbed the moderate AIS group. The other patients (n=85) showed lowered NIHSS and mRS scores and were dubbed as mild AIS. There were no significant differences between controls and either mild and moderate AIS in age, education, BMI, sex and TUD. There were no significant differences between both patient groups in the prevalence of comorbidities and the TOAST subtypes. Patients with moderate AIS showed significantly increased hsCRP levels as compared with controls and patients with mild AIS. HDL cholesterol was significantly lower in the patients as compared with controls.

**Table 1.**
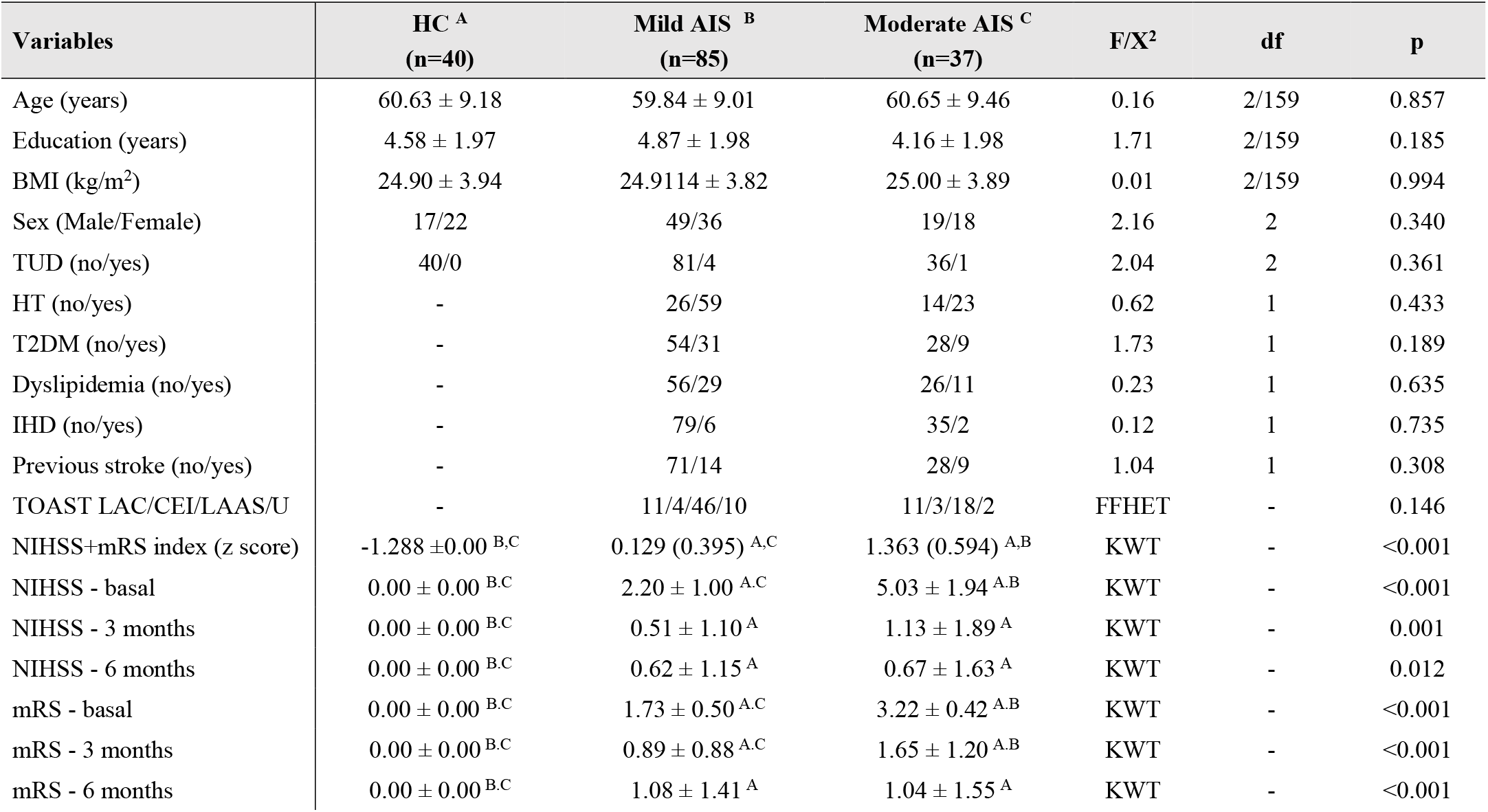
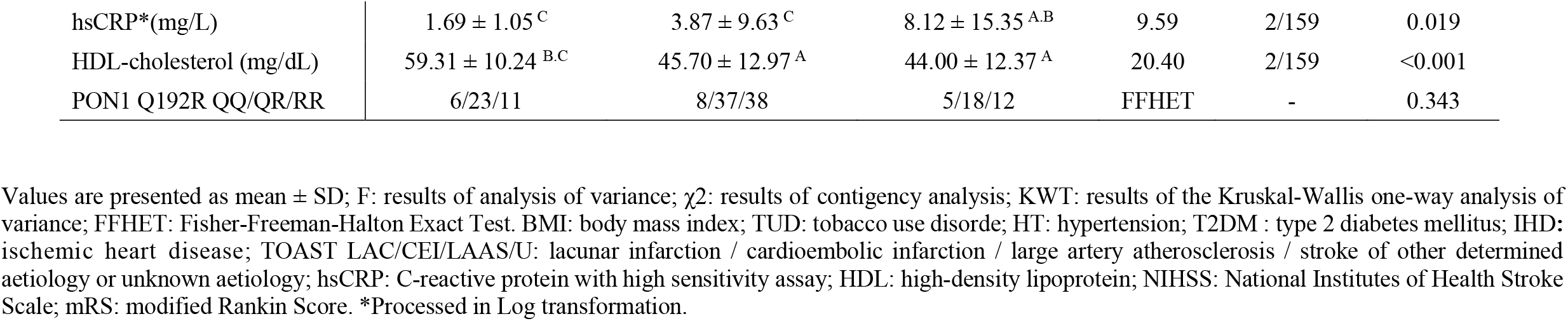
Clinical and biochemical data of healthy controls (HC) and patients with acute ischemic stroke (AIS) divided into those with mild and moderate severity.

| Variables | HC <sup>A</sup><br>(n=40) | Mild AIS <sup>B</sup><br>(n=85) | Moderate AIS <sup>C</sup><br>(n=37) | F/X<sup>2</sup> | df | p |
| --- | --- | --- | --- | --- | --- | --- |
| Age (years) | 60.63 ± 9.18 | 59.84 ± 9.01 | 60.65 ± 9.46 | 0.16 | 2/159 | 0.857 |
| Education (years) | 4.58 ± 1.97 | 4.87 ± 1.98 | 4.16 ± 1.98 | 1.71 | 2/159 | 0.185 |
| BMI (kg/m<sup>2</sup>) | 24.90 ± 3.94 | 24.9114 ± 3.82 | 25.00 ± 3.89 | 0.01 | 2/159 | 0.994 |
| Sex (Male/Female) | 17/22 | 49/36 | 19/18 | 2.16 | 2 | 0.340 |
| TUD (no/yes) | 40/0 | 81/4 | 36/1 | 2.04 | 2 | 0.361 |
| HT (no/yes) | - | 26/59 | 14/23 | 0.62 | 1 | 0.433 |
| T2DM (no/yes) | - | 54/31 | 28/9 | 1.73 | 1 | 0.189 |
| Dyslipidemia (no/yes) | - | 56/29 | 26/11 | 0.23 | 1 | 0.635 |
| IHD (no/yes) | - | 79/6 | 35/2 | 0.12 | 1 | 0.735 |
| Previous stroke (no/yes) | - | 71/14 | 28/9 | 1.04 | 1 | 0.308 |
| TOAST LAC/CEI/LAAS/U | - | 11/4/46/10 | 11/3/18/2 | FFHET | - | 0.146 |
| NIHSS+mRS index (z score) | -1.288 ± 0.00 <sup>B,C</sup> | 0.129 (0.395) <sup>A,C</sup> | 1.363 (0.594) <sup>A,B</sup> | KWT | - | <0.001 |
| NIHSS - basal | 0.00 ± 0.00 <sup>B,C</sup> | 2.20 ± 1.00 <sup>A,C</sup> | 5.03 ± 1.94 <sup>A,B</sup> | KWT | - | <0.001 |
| NIHSS - 3 months | 0.00 ± 0.00 <sup>B,C</sup> | 0.51 ± 1.10 <sup>A</sup> | 1.13 ± 1.89 <sup>A</sup> | KWT | - | 0.001 |
| NIHSS - 6 months | 0.00 ± 0.00 <sup>B,C</sup> | 0.62 ± 1.15 <sup>A</sup> | 0.67 ± 1.63 <sup>A</sup> | KWT | - | 0.012 |
| mRS - basal | 0.00 ± 0.00 <sup>B,C</sup> | 1.73 ± 0.50 <sup>A,C</sup> | 3.22 ± 0.42 <sup>A,B</sup> | KWT | - | <0.001 |
| mRS - 3 months | 0.00 ± 0.00 <sup>B,C</sup> | 0.89 ± 0.88 <sup>A,C</sup> | 1.65 ± 1.20 <sup>A,B</sup> | KWT | - | <0.001 |
| mRS - 6 months | 0.00 ± 0.00 <sup>B,C</sup> | 1.08 ± 1.41 <sup>A</sup> | 1.04 ± 1.55 <sup>A</sup> | KWT | - | <0.001 |
| hsCRP*(mg/L) | 1.69 ± 1.05<sup>C</sup> | 3.87 ± 9.63<sup>C</sup> | 8.12 ± 15.35<sup>A,B</sup> | 9.59 | 2/159 | 0.019 |
| HDL-cholesterol (mg/dL) | 59.31 ± 10.24<sup>B,C</sup> | 45.70 ± 12.97<sup>A</sup> | 44.00 ± 12.37<sup>A</sup> | 20.40 | 2/159 | <0.001 |
| PON1 Q192R QQ/QR/RR | 6/23/11 | 8/37/38 | 5/18/12 | FFHET | - | 0.343 |

Values are presented as mean ± SD; F: results of analysis of variance; $\chi^2$ : results of contingency analysis; KWT: results of the Kruskal-Wallis one-way analysis of variance; FFHET: Fisher-Freeman-Halton Exact Test. BMI: body mass index; TUD: tobacco use disorder; HT: hypertension; T2DM : type 2 diabetes mellitus; IHD: ischemic heart disease; TOAST LAC/CEI/LAAS/U: lacunar infarction / cardioembolic infarction / large artery atherosclerosis / stroke of other determined aetiology or unknown aetiology; hsCRP: C-reactive protein with high sensitivity assay; HDL: high-density lipoprotein; NIHSS: National Institutes of Health Stroke Scale; mRS: modified Rankin Score. \*Processed in Log transformation.

In the total study group, the distribution of the PON1 Q192R genotypes (QQ: n=19, QR: n=78, RR: Q=61) is consistent with Hardy-Weinberg’s equilibrium at p=0.05 (χ2=0.62, p=0.433). Table 1 shows that there is no significant association between AIS and the full PON1 genotypic model. The intercorrelation matrix showed significant partial correlations (adjusted for PON1 genotype) between HDL cholesterol and CMPAase (r=0.272, p<0.001, n=155), but not AREase activity (r=0.011, p=0.888). There was a significant partial correlation (adjusted for the genotype) between CMPAase and AREase activity (r=0.440, p<0.001), although there was no significant association between the raw PON1 activities (r=0.150, p=0.056).

### Associations between PON1 status and the diagnostic groups

**Table 2** shows the results of GLM analysis which shows the associations between AIS and the different indices of basal PON1 activity. There was a strong association between the PON1 activity indices and the diagnosis (AIS versus controls) and the PON1 Q192R genotype. Table 2 shows the results of the univariate GLM for the between-subject effects whilst **Table 3** shows the model-generated estimated marginal mean values of the PON1 indices (after controlling for the effects of the Q192R gene variants) in AIS versus controls indicating that all indices used here were significantly different between both groups. The top-2 strongest effects were estblished for zCMPAase-zAREase and zCMPAase+zHDL. While basal CMPAase was decreased in AIS patients with a low effect size (0.032), AREase activity was significantly increased with a much higher effect size (0.111). Both the zCMPAase-zAREase and zCMPAase+zHDL indices were significantly lower in stroke patients than in controls. We have rerun the same analyses without controlling for the effects of the PON1 genotype and these analyses yielded similar resuls.

**Table 2.** Results of general linear models (GLM) which show the associations between acute ischemic stroke (AIS) and different indices of paraoxonase activites

| Type | Dependent variable | Explanatory variable | F | df | p | Partial Eta Squared |
| --- | --- | --- | --- | --- | --- | --- |
| Univariate GLM | CMPAase | AIS versus HC | 5.02 | 1/154 | 0.026 | 0.032 |
|  | AREase |  | 19.16 | 1/154 | <0.001 | 0.111 |
|  | zCMPAase-zAREase |  | 51.79 | 1/154 | <0.001 | 0.252 |
|  | zCMPAase+zHDL |  | 33.54 | 1/154 | <0.001 | 0.179 |
|  | CMPAase | PON1 Q192R genotype | 19.64 | 2/154 | <0.001 | 0.203 |
|  | AREase |  | 22.57 | 2/154 | <0.001 | 0.227 |
|  | zCMPAase-zPON |  | 95.49 | 2/154 | <0.001 | 0.554 |
|  | zCMPAase+zHDL |  | 9.14 | 2/154 | <0.001 | 0.106 |
|  | CMPAase Month 3 | AIS versus HC | 21.14 | 1/111 | <0.001 | 0.160 |
|  | AREase Month 3 |  | 14.47 | 1/111 | <0.001 | 0.115 |

HC: healthy controls; CMPAase: 4-(chloromethyl)phenyl acetate hydrolysis; PON1: Paraoxonase; AREase: arylesterase; HDL: high density lipoprotein. All biomarkers are measured in baseline conditions, except when stated otherwise.

**Table 3.** Model-generated estimated marginal mean values of different paraoxonase indices in patients with acute ischemic stroke (AIS) versus healthy controls (HC) and between the PON1 Q192R geneotypes.

| Variables | Controls<br>N=40 | AIS<br>N=118 |  |
| --- | --- | --- | --- |
| CMPAase (U/mL) | 38.38 ± 1.56 | 34.50 ± 1.00 |  |
| AREase (U/mL) | 254.01 ± 16.53 | 334.44 ± 10.66 |  |
| zCMPAase-zAREase (z scores) | 0.320 ± 0.107 | -0.538 ± 0.069 |  |
| zCMPAase+zHDL (z scores) | 0.609 ± 0.148 | -0.342 ± 0.095 |  |
| CMPAase Month 3 (U/mL) | 38.19 ± 1.55 | 30.35 ± 1.11 |  |
| AREase Month 3 (U/mL) | 244.27 ± 10.45 | 197.68 ± 7.95 |  |
| Variables | PON1 Q192R QQ<sup>A</sup><br>n=19 | PON1 Q192R QR<sup>B</sup><br>n=78 | PON1 Q192R RR<sup>C</sup><br>n=61 |
| CMPAase (U/mL) | 29.55 ± 2.17<sup>B,C</sup> | 36.18 ± 1.12<sup>A,C</sup> | 43.58 ± 1.32<sup>A,B</sup> |
| AREase (U/mL) | 384.26 ± 23.08<sup>A,C</sup> | 284.175 ± 11088<sup>A,C</sup> | 214.23 ± 14.03<sup>A,B</sup> |
| zCMPAase-zAREase (z scores) | -1.278 ± 0.150<sup>B,C</sup> | -0.015 ± 0.077<sup>A,C</sup> | 0.965 ± 0.091<sup>A,B</sup> |
| zAREase+zHDL (z scores) | -0.252 ± 0.206<sup>B,C</sup> | 0.059 ± 0.106<sup>A,C</sup> | 0.593 ± 0.125<sup>A,B</sup> |
| CMPAase Month 3 (U/mL) | 27.24 ± 2.18<sup>B,C</sup> | 34.58 ± 1.14<sup>A,C</sup> | 40.99 ± 1.43<sup>A,B</sup> |
| AREase Month 3 (U/mL) | 260.48 ± 21.77<sup>C</sup> | 222.32 ± 8.16<sup>C</sup> | 180.13 ± 10.32<sup>A,B</sup> |

CMPAase: 4-(chloromethyl)phenyl acetate hydrolysis; AREase: arylesterase; HDL: high density lipoprotein. All biomarkers are measured in baseline conditions, except when stated otherwise.

Table 2 and Table 3 show that CMPAase (0.203) and AREase (0.227) were both partly determined by the genotypes, with lower CMPAase but higher AREase activity in the QQ genotype than the RR genotype. The zCMPAase-zAREse and zCMPAase+zHDL ratios were signicantly different between the three genotypes and increased from the QQ to the QR to the RR genotype.

### Prediction of AIS versus controls

The association between PON1 status and AIS was examined using neural network analysis. We entered 13 variables namely PON1 genotypes, CMPAase, AREase, zCMPAase-zAREase, zCMPAase+zHDL, hypertension, previous stroke, dyslipidemia, BMI, age and sex. The feedforward network was trained using two hidden layers, the first with four units and the second with three. In the hidden layers, we utilized hyperbolic tangent as the activation function, and in the output layer, we used identity. The sum of squares error term in the testing (2.454) set was lower than in the training (3.095) set, demonstrating that the model has learnt to generalize from the trend. The proportion of inaccurate classifications remained relatively similar throughout the training (4.5%), testing (4.5%), and holdout (6.5%) samples, demonstrating that the model is not overtrained and sufficiently replicable. The AUC ROC curve was 0.975, and the sensitivity and specificity in the holdout sample were 96.7% and 87.5%, respectively. The (relative) relevance of the input factors in predicting stroke vs controls is shown in **Figure 1**. The top-6 most important variables with the greatest predictive power of the model were zCMPAase-zAREase and hypertension, followed at a distance by AREase, dyslipidemia, zCMPAase+zHDL, prior stroke, BMI, and PON1 RR and QR genotypes, whilst the other variables were less important.

**Figure 1.**
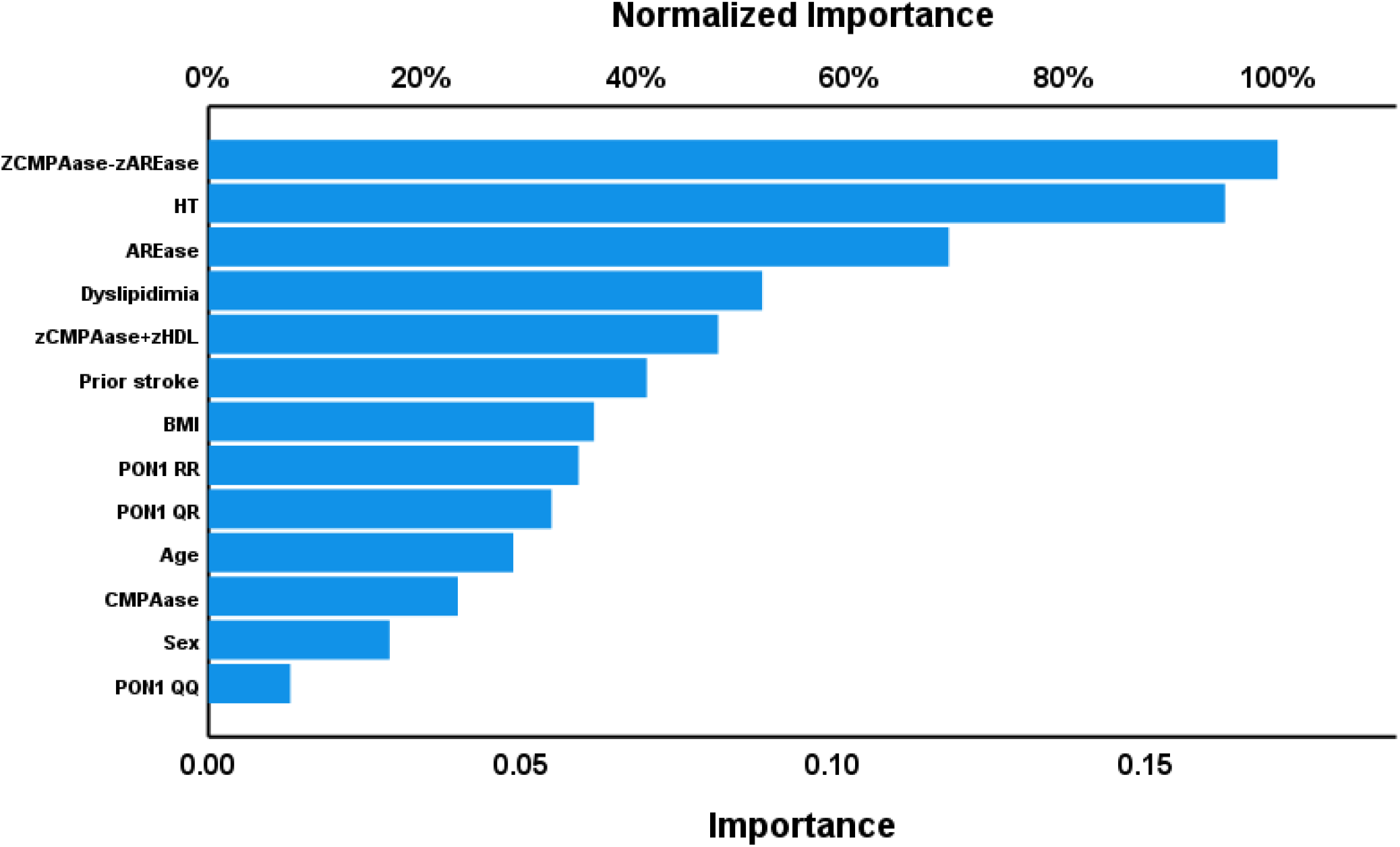
Results of neural network analysis showing the relevance chart. This bargraph shows the (relative) relevance of the explanatory variables predicting stroke vs controls. The top-4 most important variables with the greatest predictive power of the model were zCMPAase+zHDL, hypertension, HDL cholesterol and AREase, which were followed at a distance by previous stroke, PON1 RR genotype,, and again at a distance by dyslipidemia and zResCMPAase- zResAREase, whilst the other variables were less important.

Logistic regression analysis showed that zResCMPAase-zResAREase (W=23.17, df=1, p<0.001, Odds-ratio=0.262, 95% CI: 0.152 and 0.452) and HDL (W=18.87, df=1, p<0.001, Odds-ratio=0.273, 95% CI: 0.152 and 0.491) predicted stroke with an accuracy of 86.7%, sensitivity of 84.7% and specificity of 92.5%. The Nagelkerke pseudo R^2^ effect size was 0.542.

### Effects of time on PON1 activities

**Table 4** shows the results of GEE, repeated measures, performed in patients only, with both PON1 activities as repeated measures (from baseline to month 3) and the effects of time, PON1 genotype and the time X genotype interaction as explanatoy variables. We found that CMPAase activity decreased significantly from baseline to 3 month later. In addition, also the AREase activity decreased from baseline to 3 months later and the significant time X genotype interaction showed that this effects was most pronounced in the QQ genotype.

**Table 4.** Results of Generalized Estimating Equations (GEE), repeated measures, showing the effects of time (differences between levels at baseline and month 3) on 4-(chloromethyl)phenyl acetate hydrolysis (CMPAase) and arylesterase (AREase) activities.

| Dependent variables | Explanatory variables | Baseline (mean SE) | Month 3 (mean, SE) | Wald/$\chi^2$ | df | P |
| --- | --- | --- | --- | --- | --- | --- |
| CMPAase from baseline month 3 (U/mL) | Time | 34.59 (0.88) | 30.40 (0.87) | 27.22 | 1 | <0.001 |
|  | PON1 Q192R genotype |  |  | 51.55 | 2 | <0.001 |
| AREase from baseline to month (U/mL) | Time | 340.78 (11.82) | 197.72 (8.49) | 115.24 | 1 | <0.001 |
|  | PON1 Q192R genotype |  |  | 31.53 | 2 | <0.001 |
|  | Time X Q192R genotype |  |  | 13.01 | 2 | 0.001 |
|  | PON1 Q192R QQ | 444.92 (27.85) | 230.29 (20.01) |  |  |  |
|  | PON1 Q192 QR | 319.66 (16.20) | 196.77 (9.69) |  |  |  |
|  | PON1 Q192R RR | 257.76 (148.53) | 169.11 (14.00) |  |  |  |

HC: healthy controls; CMPAase: 4-(chloromethyl)phenyl acetate hydrolysis; PON1: Paraoxonase; AREase: arylesterase; HDL: high density lipoprotein.

### Prediction of basal NIHSS and mRS scores

**Table 5** shows the results of multiple regression analyses with the NIHSS and mRS scores as depedent variables and the PON1 status data as explanatory variables whilst we allowed for the effects of confounders (including comorbidities and hsCRP, BMI, age and sex). Around 28.0% of the variance in the baseline NIHSS score could be explained by the regression on hypertension, ResAREase (both positively associated) and ResCMPAase (inversely associated) (see model#1). Model #2 demonstrates that substituting CMPAase and AREase for their composite score produced equivalent results. The best prediction (model#3) was obtained by adding HDL cholesterol which increased the explaned variance by 6.7%. **Figure 2** shows the partial regression of the baseline NIHSS score on the zResCMPAase-zResAREase score.

**Table 5.** Results of multiple regression analysis with National Institutes of Health Stroke (NIHSS) and Modified Rankin Score (mRS) scores as depedent variables.

| Dependent variable | Explanatory variable | $\beta$ | t | p | F<sub>model</sub> | df | p | R<sup>2</sup> |
| --- | --- | --- | --- | --- | --- | --- | --- | --- |
| NIHSS Basal | **Model #1** |  |  |  | 19.84 | 3/153 | <0.001 | 0.280 |
|  | HT | 0.301 | 4.20 | <0.001 |  |  |  |  |
|  | Res AREase | 0.356 | 4.46 | <0.001 |  |  |  |  |
|  | Res CMPAase | -0.320 | -4.07 | <0.001 |  |  |  |  |
|  | **Model #2** |  |  |  | 29.82 | 2/154 | <0.001 | 0.279 |
|  | zResCMPAase– zResAREase | -0.354 | -4.97 | <0.001 |  |  |  |  |
|  | HT | 0.308 | 4.25 | <0.001 |  |  |  |  |
|  | **Model #3** |  |  |  | 20.34 | 4/153 | <0.001 | 0.347 |
|  | HT | 0.235 | 3.32 | 0.001 |  |  |  |  |
|  | HDLcholesterol | -0.274 | -3.86 | <0.001 |  |  |  |  |
|  | Res AREase | 0.337 | 4.43 | <0.001 |  |  |  |  |
|  | Res CMPAase | -0.241 | -3.12 | 0.002 |  |  |  |  |

HT: hypertension; NIHSS: National Institutes of Health Stroke Scale; mRS: modified Rankin Score. CMPAase: 4-(chloromethyl)phenyl acetate hydrolysis; PON1: Paraoxonase; AREase: arylesterase.

**Figure 2.**
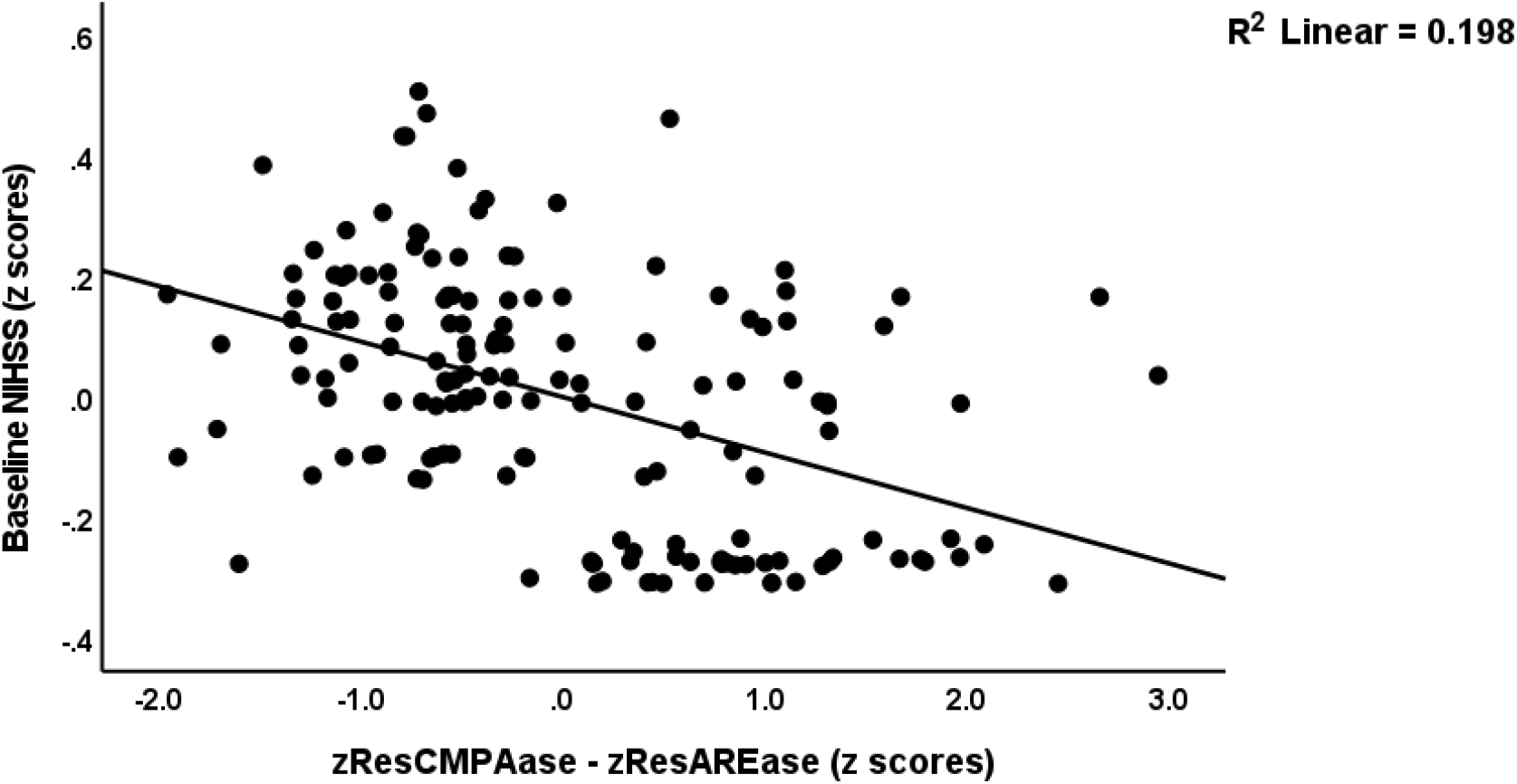
shows the partial regression of the baseline NIHSS score on the zResCMPAase-zResAREase score.

**Table 6** displays the results of various regression models using the baseline mRS scores as dependent variables, as well as the mRS values at months 3 and 6. The baseline mRS score was best predicted (30.9% of the variance) by hypetension (positively), zResCMPAase-zResAREase and zCMPASe+zHDL (both inversely associated) (model#1). The zResCMPAase-zResAREase score was inversely associated with baseline mRS and explained 17.4% of its variance, whilst this score together with previous stroke explained 20.6% (see model#2). **Figure 3** shows the partial regression of baseline mRS score on the zCMPAase + zHDL score (after controlling for age, sex, BMI and genotypes). The mRS score at 3 months was best predicted by hypertension and the zResCMPAase-zResAREase score (see model#3). The mRS score at 6 months was best predicted by dyslipidemia, the zResCMPAase-zResAREase score and previous stroke which together explained 19.6% of the variance (see model#4).

**Table 6.**
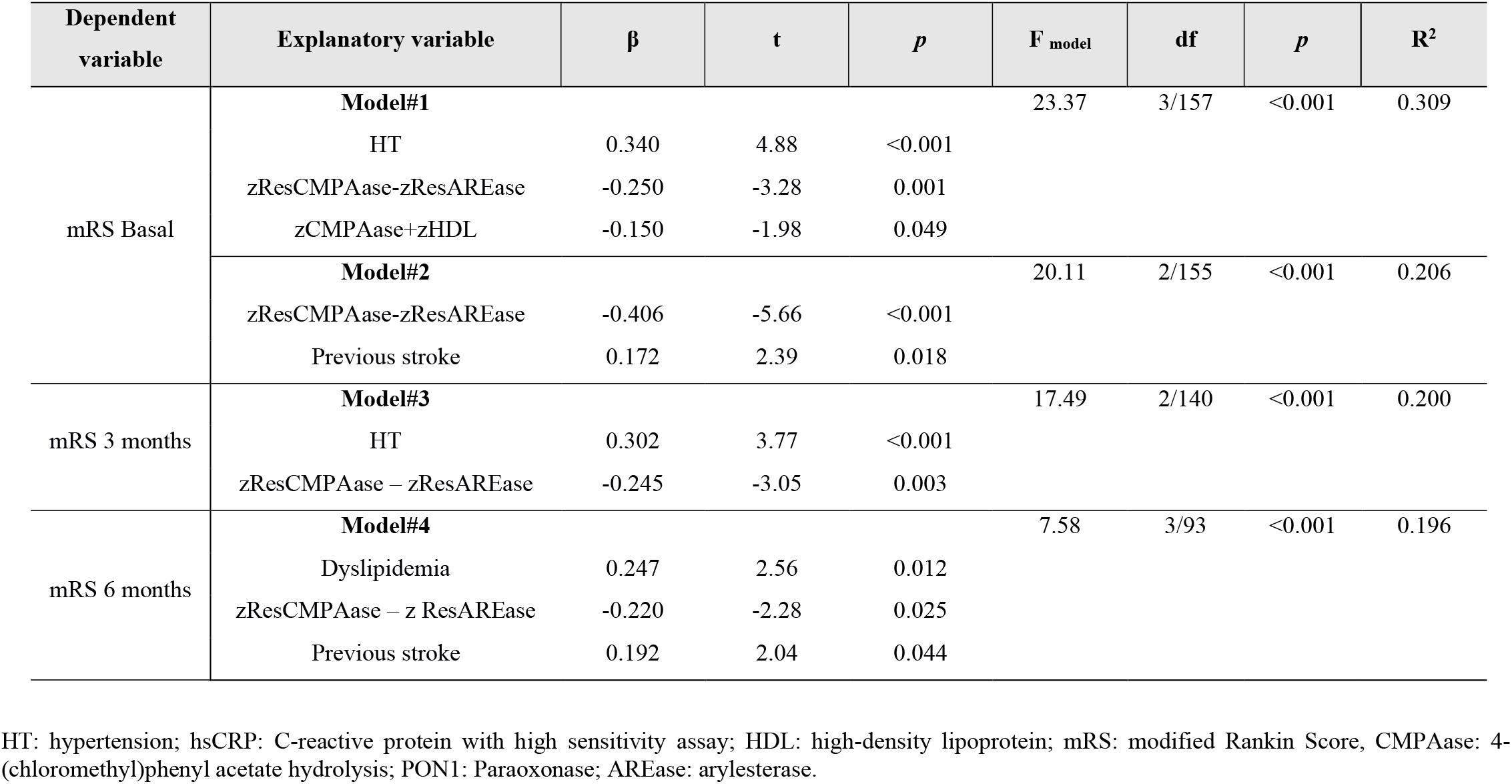
Results of multiple regression analyses with Modified Rankin Score (mRS) scores as depedent variables.

**Figure 3.**
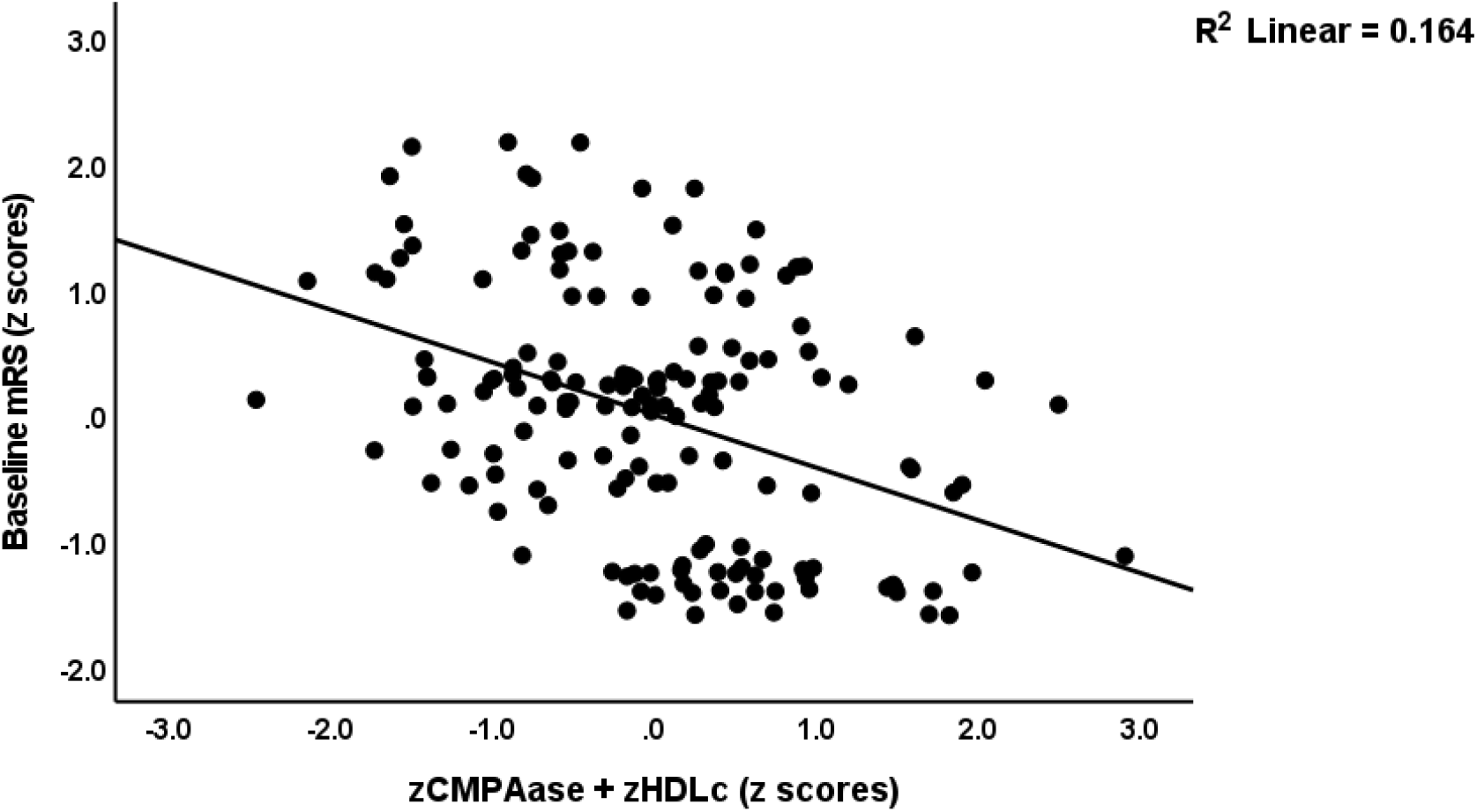
shows the partial regression of baseline mRS score on the zCMPAase + zHDL score (after controlling for age, sex, BMI and genotypes).

### Results of GEE performed on the repeated measures of the NIHSS and mRS scores

**Table 7** shows the results of GEE with the repeated measurements of the NIHSS and mRS scores as dependent variables and PON1 status and HDL as explanatory variables. Different repeated measures were entered, namely the NIHSS/mRS scores from baseline to month 3, from month 3 to month 6, and from baseline to month 3 to month 6. Baseline PON1 measurements as well as the repeated PON1 measurement from basal to month 3 were entered as explanatory variables. In all cases we found that one of the CMPAase indices was inversely associated with the disability outcomes, whilst one of the AREase indices was positively associated with the disability scores (except for NIHSS from month 3 to 6). In addition, the PON1 genotypes and HDL significantly impacted most disability data.

**Table 7.** Results of Generalized Estimating Equations (GEE), repeated measures, with the National Institutes of Health Stroke (NIHSS) and Modified Rankin Score (mRS) scores as dependent variables and Paraoxonase (PON)1 status as explanatory variable.

| Dependent variables | Explanatory variables | B | SE | Lower 95% CI | Upper 95% CI | Wald/X2 (df=1) | p |
| --- | --- | --- | --- | --- | --- | --- | --- |
| NIHSS M0–M3 | z CMPAase – z AREase M0-M3 | -0.552 | 0.1343 | -0.815 | -0.288 | 145.27 | <0.001 |
| NIHSS M0-M3 | PON1 Q192R QQ | -0.778 | 0.2841 | -0.910 | -0.221 | 7.51 | 0.016 |
|  | Res CMPAase M0-M3 | -0.641 | 0.1690 | -0.972 | -0.310 | 14.39 | 0.028 |
|  | Res AREase M0-M3 | 1.073 | 0.1877 | 0.705 | 0.705 | 32.69 | 0.019 |
| NIHSS M3-M6 | PON1 Q192R additive | 0.411 | 0.1702 | 0.077 | 0.744 | 5.82 | 0.016 |
|  | HDL baseline | -0.012 | 0.0055 | -0.023 | -0.001 | 4.83 | 0.028 |
|  | CMPAase M0-M3 | -0.289 | 0.1231 | 0.530 | -0.047 | 5.50 | 0.019 |
| NIHSS M0-M3-M6 | PON1 Q192R additive | 0.887 | 0.1735 | 0.547 | 0.921 | 26.12 | 0.045 |
|  | zCMPAase–zAREase baseline | -0.470 | 0.0910 | -0.648 | -0.292 | 26.72 | <0.001 |
|  | HDL baseline | -0.026 | 0.0059 | -0.038 | -0.015 | 19.59 | <0.001 |
| mRS M0-M3 | PON1 Q192R QQ | -0.468 | 0.2360 | -0.930 | -0.005 | 3.93 | 0.047 |
|  | Res CMPAase M0-M3 | -0.344 | 0.0936 | -0.528 | -0.161 | 13.52 | <0.001 |
|  | Res AREase M0-M3 | 0.524 | 0.0823 | 0.363 | 0.685 | 40.51 | <0.001 |
| mRS M3-M6 | PON1 Q192R additive | 0.295 | 0.1394 | 0.021 | 0.568 | 4.47 | 0.035 |
|  | zCMPAase-zAREare M0-M3 | -0.173 | 0.0846 | -0.339 | -0.007 | 4.18 | 0.041 |
|  | HDL baseline | -0.019 | 0.0049 | -0.028 | -0.009 | 14.83 | <0.001 |
| mRS M0-M3-M6 | PON1 Q192R additive | 0.585 | 0.1152 | 0.360 | 0.811 | 25.80 | <0.001 |
|  | zCMPAase-zAREase baseline | -0.326 | 0.0632 | -0.450 | -0.202 | 26.60 | <0.001 |
|  | HDL baseline | -0.021 | 0.0043 | -0.029 | -0.012 | 22.56 | <0.001 |

CMPAase: 4-(chloromethyl)phenyl acetate hydrolysis; PON1: Paraoxonase; AREase: arylesterase; HDL: high density lipoprotein.
M0-M3: assessments at baseline (M0) and month 3 (M3) were entered as two repeated measurements
M3-M6: assessments at baseline (M0) and month 3 (M3) were entered as two repeated measurements
M0-M3-M6: assessments at baseline (M0), month 3 (M3) and month 6 (M6) were entered as three repeated measurements

### Results of PLS analysis

**Figure 4** shows the final PLS path model. depicts the final PLS model, which only displays significant paths and indicators. All variables were entered as single indicators except the phenome of stroke, which was conceptualized as a latent vector extracted from the baseline NIHSS and mRS scores and the ordinal grouping as controls, and mild and moderate AIS. Our model assumes that the effects of the PON1 gene on the phenome are mediated by PON1 activities, although we also allow for direct effects. CRP and hypertension are considered to impact both PON1 activities as well as HDL while the zCMPAase+zHDL composite reflect the antioxidant complex between both factors. We found adequate convergence and construct reliability validity values for the baseline latent vector with AVE = 0.942, Cronbach alpha = 0.969, rho A= 0.970, and composite reliability = 0.980) while all outer loadings were > 0.961 at p<0.0001. Blindfolding revealed acceptable construct redundancies for the latent vector of 0.358, whilst CTA showed that the latent vectors was not specified incorrectly as a reflective model. With an SRMR of 0.011, the overall quality of the model is more than adequate. According to PLSPredict, all construct indicator Q2 predict values were positive, indicating that the prediction error was less than the most conservative benchmark. We found that 39.3% of the variance in the stroke phenome was explained by AREase, hypertension, PON1 gene (additive model was most adequate), CRP (all positively) and zCMPAase+zHDL (inversely). Both CRP and hypertension had significant direct (see Figure 4) and indirect effects on the phenome yielding significant total effects (t=3.55, p<0.001, and t=5.53, p<0.001, respectively). The PON1 genotype had significant direct effects on the phenome but also indirect effects that were mediated via CMPAase (t=-4.78, p<0.001) and AREase, yielding a total effect that is not significant (t=1.00, p=0.159).

**Figure 4.**
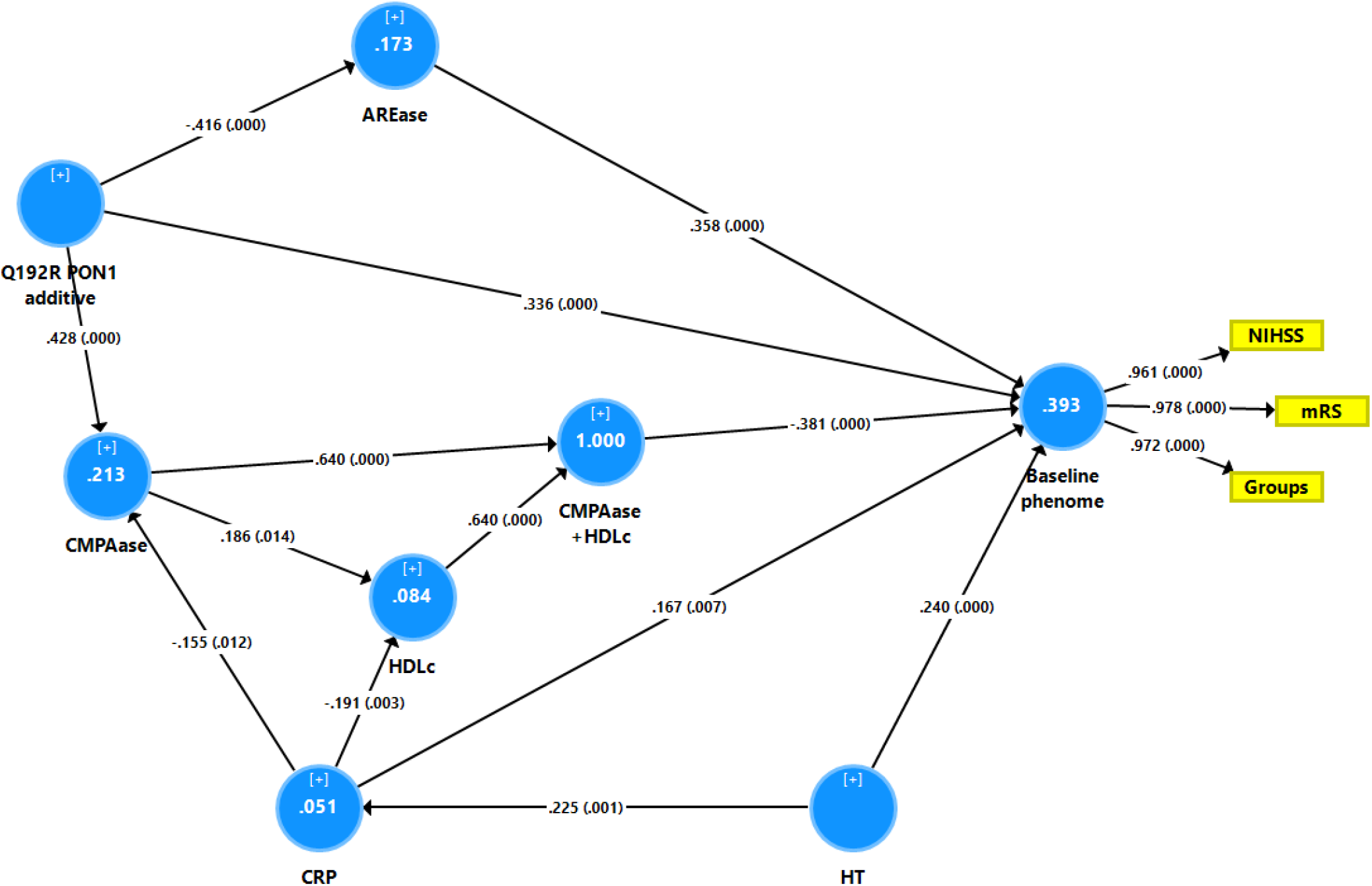
shows the final PLS path model. depicts the final PLS model, which only displays significant paths and indicators. All variables were entered as single indicators except the phenome of stroke, which was conceptualized as a latent vector extracted from the baseline NIHSS and mRS scores and the ordinal grouping as controls, and mild and moderate AIS. Our model assumes that the effects of the PON1 gene on the phenome are mediated by PON1 activities, although we also allow for direct effects. CRP and hypertension are considered to impact both PON1 activities as well as HDL while the zCMPAase+zHDL composite reflect the antioxidant complex between both factors.

## Discussion

### CMPAase and AREase activities in AIS

The first major finding of the present study is that increased AREase and reduced CMPAase activities are associated with AIS, and that these enzyme activities highly predict the degree of post-stroke impairments (mRS and NIHSS scores) at baseline and 3 and 6 months after AIS. Interestingly, both CMPAase and AREase activity further decreased from baseline to 3 months later.

PON1 is capable of hydrolyzing many lactones, arylesters, organophosphate insecticides, nerve gases, glucuronide medicines, and estrogen esters (Rajkovic et al. 2011). Using different substrates, the activity of PON1 can be evaluated to determine paraoxonase/phosphotriesterase activity (using paraoxon or CMPA as substrates), AREase activity (using phenylacetate or 4 (p) -nitrophenyl acetate as substrates), or lactonase activity (5-thiobutyl butyrolactone or dihydrocoumarin as substrates) (Ceron et al., 2014).

Our work is the first to our knowledge to establish a correlation between PON1 CPMAase activity and AIS. Previous studies have shown a negative correlation between overall PON 1 activity and AIS (Ozturk et al., 2020; Xu et al., 2021). In some investigations, an inverse correlation between AREase serum activity and AIS was identified (Shenhar-Tsarfaty et al., 2013; Castellazzi et al., 2016). Some studies discovered that serum PON1 activity is connected with the functional prognosis of AIS patients and indicated that the mRS score of AIS patients tends to decrease as serum PON1 activity levels rise (Shenhar-Tsarfaty et al., 2013; Xu et al., 2021). On the other hand, Abdullah et al. (2013) observed no association between serum PON1 activity and NIHSS in AIS patients. However, it is difficult to compare these findings to ours since earlier research overlooked the diverse enzyme activities of PON1. In this respect, CMPAase activity is also inversely associated with other neurodegenerative and neuroinflammatory disorders, such as deficit schizophrenia (Matsumoto et al., 2021), temporal lobe epilepsy (Michelin et al., 2022), and mood disorders, such as major depression and bipolar disorder (Moreira et al., 2019b), as well as the severity, progression, and neurocognitive impairments that accompany these disorders. Nevertheless, according to our findings, AIS is the only condition accompanied by concurrent increases in AREase and reductions in CMPAase activity.

Our findings that CMPAase (decreased) and AREase (increased) are differently associated with AIS and subsequent impairments demonstrate once again that the PON1 enzyme has (at least) two unique catalytic sites (CMPAase and AREase) (Moreira et al., 2019a). A newly estimated z unit-based composite score based on decreased CMPAase vs increased AREase activity was the best predictor of AIS and the mRS and NIHSS scores 3 and 6 months after acute stroke in our research. This demonstrates that decreased CMPAase coupled with elevated AREase are key contributors to AIS and subsequent disabilities. It appears that the PON1 enzyme has a Janus-face with two distinct catalytic sites that are differentially associated with AIS. As a result, we recommend assessing both CMPAase and AREase activities and computing their z-unit composite score as indicators of PON1 activity.

### PON1 activities, HDL and AIS

The second major finding of our study is that CMPAase, but not AREase, activity was significantly correlated with HDL levels and that lowered levels of the newly calculated composite score of zCMPAase+zHDL (or combinations of low HDL with low CMPAase in GEE models) were the second-best predictors of AIS and its outcome. After its release in serum, PON1 binds to HDL thereby contributing to protective functions of HDL and preventing oxidative alterations of HDL and LDL, which are linked to several vascular disorders, including atherosclerosis (Kim et al., 2007; Furtado et al., 2018; Moreira et al., 2019a). In fact, the PON1 enzyme accounts for the majority of HDL’s antioxidant properties and shows anti-inflammatory, anti-apoptotic and antiatherogenic properties (Marsillach et al., 2021; Moreira et al., 2019a).

Most AIS are thromboembolic in origin, originating from an atherosclerotic plaque (Campbell et al., 2019), whilst reduced HDL and increased triglycerides and LDL are important biomarkers and risk factors of AIS because they are involved in the process of atherosclerosis (Benjamin et al. 2018; Norrving et al., 2018; Kuriakose et al., 2020). Several mechanisms have been proposed for the role of PON1 in the atheroprotective activity of HDL, including: a) making HDL more resistant to oxidation, thereby preserving its antiatherogenic capacity; b) inducing reverse cholesterol transport; c) decreasing and preventing oxidation of LDL by reducing the formation of foam cells; d) attenuated macrophage production of reactive oxygen radicals and inhibiting monocyte chemoattractant protein-1 (MCP) (Marchegiani et al., 2008; Shah, 2010, Tajbakhsh et al., 2017; Tisato et al., 2019). Moreover, the PON1-HDL complex has antioxidant, anti-inflammatory, antiapoptotic, vasorelaxant, and antithrombotic characteristics and supports endothelial function normalization and endothelial progenitor cell function stimulation (Shah, 2010; Campbell et al., 2019; Moreira et al., 2019a).

The newly calculated composite score of zCMPAase+zHDL, therefore, represents the activity and functionality of the PON1-HDL complex in serum (Maes et al., 2018a; 2018b). Consequently, our data revealing that this composite score is much lower in AIS and is inversely associated with post-stroke disabilities is in agreement with the knowledge that decreased antioxidant, anti-inflammatory, anti-apoptotic, and atherogenic protection is a crucial component of the pathophysiology of AIS. The loss of the above protective mechanisms (lowered CMPAase) may potentially lead to increased production of reactive oxygen species and activation of oxidative and inflammatory pathways, thereby interfering with the ability to protect HDL and LDL from oxidation (Varela et al., 2020; Moreira et al., 2019a; Kotur-Stevuljevic et al., 2015; Tajbakhsh et al., 2017).

It should be emphasized that serum CMPAase activity (but not AREase activity) was substantially linked with serum HDL concentrations in some neuropsychiatric illnesses (bipolar disorder and major depression) (Maes et al., 2018a; 2018b). Moreover, mediation analysis demonstrated that the effects of CMPAase or the CMPAase-HDL complex on the phenome of these mood disorders, schizophrenia, and epilepsy are mediated by the effects of lipid peroxidation, including increased malondialdehyde formation (Maes et al., 2022; Maes et al., 2018b; Maes et al., 2020). These data may suggest that CMPAase in contrast to AREase is associated with increased oxidative stress and, as a result, exhibits higher antioxidant qualities than AREase, at least in neuropsychiatric disorders and AIS. Diabetes mellitus, rheumatoid arthritis, systemic lupus erythematosus, liver and kidney disorders, including renal failure, psoriasis, and macular degeneration, are other diseases with a significant inflammatory and /or oxidative component that have been linked to dysfunctional PON1 and/or HDL (Goswami et al., 2009; Moreira et al., 2019a). Future study should explore the two catalytic PON1 sites in those disorders and delineate the differences in substrates and properties of these different sites.

### Inflammation, hypertension and PON1 activities in AIS

The third major finding of our study is that CRP influenced CMPAase (but not AREase) activity, HDL, and the CMPAase-HDL association (see results of PLS analysis). A recent meta-analysis showed that increased baseline hsCRP is associated with increased risk of AIS (Zhou et al., 2016). Alfieri et al. (2020) found that hsCRP is highly linked with AIS and consequent disabilities at baseline and 3 months later. Moreover, higher CRP levels are related with poor outcomes as measured by the NIHSS scale, stroke subtypes, and traditional risk factors (Matsuo et al., 2016) and are reported as independent predictors of 1-year mortality (Li, Liu, 2015). CRP is thus a significant inflammatory biomarker of AIS and post-stroke prognosis (Bustamante et al., 2016; Bustamante et al., 2017; Geng et al., 2016; Yu et al., 2017). Low PON1 activity has been associated with elevated CRP levels, indicating a mechanistic connection between PON1 activity and inflammation and the development of atherosclerosis (Bhattacharyya et al., 2008).

Our findings also show that these decreases in CMPAase activity and in the zCMPAase-zAREase score from baseline to three months later predicted increased disability scores from baseline to 3 and 6 months later. It is plausible that the lowered CMPAase and AREase activities from baseline to three months later are caused by the above inflammatory processes. Activated macrophages promote the release of peroxides, nitric oxide, and myeloperoxidase, which, when combined, may form peroxynitrite and hypochlorous acid (Maes et al., 2018a). The latter are toxic products that may damage PON1 in HDL, whilst myeloperoxidase may further oxidize PON1, inactivating PON1 and reducing its binding to HDL (Menini, Gugliucci, 2014). Moreover, during systemic inflammation, native HDL may be transformed to dysfunctional HDL owing to the enzyme PON1’s loss of antiatherogenic capabilities, which leads to the less protective and more atherogenic character of HDL (Nessler et al., 2018).

Our investigation also revealed that the effects of hypertension on AIS are partly mediated by the effects of CRP on CMPAase and the CMPAase-HDL complex. The most frequently reported risk factor for AIS is hypertension (Laaksonen et al., 2008; Tohidi et al., 2012; Wajngarten, Silva., 2019). Inflammation, oxidative stress, and endothelial dysfunction have been linked to both hypertension and AIS pathogenesis (Kunutsor et al., 2017). Low PON-1 activity in the circulation has been observed in hypertensive individuals (Uzun et al., 2004; Yildiz et al., 2008) and HDL may protect against hypertension due to its antithrombotic, antioxidant and anti-inflammatory effects, which reduce damage to the blood vasculature. HDL is also antiatherogenic due to its well-known role in reverse cholesterol transport, which modulates endothelial function in a favorable manner (Sitia et al., 2010). In addition, the underlying mechanisms through which HDL may reduce the incidence of hypertension comprise antioxidant and anti-inflammatory capabilities, which in fact are due to its PON1 binding (Kaypakl et al., 2016, Demir et al., 2019).

### PON1 genotypes and AIS

Although there was no direct association between PON1 Q192R genotypes and AIS, the influence of PON1 genotypes on stroke became evident when multiple regression, GEE and PLS analyses were conducted. These analyses demonstrated a protective impact of QQ on NIHSS scores 3 and 6 months after baseline and that the RR genotype was linked with elevated mRS values 3 and 6 months after AIS. Liu et al. (2013) conducted a comprehensive review and meta-analysis of AIS and PON1 gene polymorphisms and determined that the R allele or RR genotype of the PON1 Q192R polymorphism was associated with an elevated risk of AIS in the general population (Liu et al., 2013). The homozygous QQ genotype may be protective against AIS in a Turkish population (Demirdöğen et al., 2009). Nonetheless, not all investigations have confirmed these results (Huang et al., 2006; Xiao et al., 2009). One possibility is that the distribution of the PON1 genotype varies greatly across ethnic groups (Koda et al., 2004; Richter et al., 2010; Gupta et al., 2022) and that the altered PON1 genetic variations among different ethnicities may influence their vulnerability to AIS (Tajbakhsh et al., 2017). The benefits of the PON1 Q192R genotype on AIS have been attributed to the Q isoform inhibiting LDL oxidation more effectively than the R isoform (Kim et al., 2012; Menini, Gugliucci, 2014). In addition, the R isoenzyme has less ability to hydrolyze lipid peroxide and, thus, poorer anti-atherogenic properties than the 192Q isoenzyme (Liu et al., 2010; Zhao et al., 2019). Nevertheless, the R form has certain benefits since it can hydrolyze paraoxon more quickly than the Q type (Dounousi et al., 2016).

Previous research showed that patients with the PON1 QQ genotype demonstrate reduced PON1 activity at baseline and 12 months after a stroke compared to those with the RQ/RR genotypes (Shenhar-Tsarfaty et al., 2013). However, our data also indicate that the PON1 QQ genotype, which may have protective effects against AIS, substantially decreases CMPAase and increases AREase activity, which both increase risk to AIS. Furthermore, our PLS analysis revealed that the associations between the PON1 Q192R genotype, the PON1 activities and AIS are far more complex than can be estimated from association studies or regression analysis. Thus, our PLS analysis demonstrates that a) the RR genotype is associated with increased AIS risk via a direct pathway, b) the QQ genotype is associated with increased AIS risk via two indirect pathways, namely via CMPAase (and the CMPAase-HDL complex) and AREase, which both yield a negative direction despite the fact that the effects of the genotype on both enzymatic activities are completely opposite; and c) the direct and indirect paths have opposite effects leading to an overall effects that is not significant. Thus, the PON1 gene has complex, multiple Janus-faced effects on AIS outcome which can only be delineated using mediation analysis and including both catalytic enzyme activities. This view is supported by our neural network results which revealed that the Q192R genotype and both PON1 activities contribute considerably to the prediction of AIS.

## Conclusions

According to our results, the PON1 enzyme contains (at least) two distinct catalytic sites, CMPAase and AREase activity. AIS is accompanied by simultaneous decreases in CMPAase and increases in AREase activity. zCMPAase+zHDL is another major predictor of AIS and AIS-related impairments, highlighting the significance of the CMPAase-HDL complex for the oxidative, inflammatory, and atherogenic pathophysiology underlying AIS. The PON1 Q192R genotype has strong, although complicated, impacts on AIS outcome and disability. We propose that future AIS research evaluate the PON1 status, which should include the PON1 Q192R genotype, CMPAase, and AREase activity, and should be analysed utilizing multistep mediation analysis. The data indicate that both catalytic PON1 sites and the CMPAase-HDL complex are relevant targets for the prevention and treatment of AIS, as well as for the attenuation or prevention of AIS-related disabilities.

## Data Availability

All data produced in the present study are available upon reasonable request to the authors

## Author Declarations

### Availability of data and materials

The dataset generated during and/or analyzed during the current study will be available from MM upon reasonable request and once the authors have fully exploited the dataset.

### Author’s Contributions

All authors contributed to the paper. MM and CT designed the study. Statistical analyses were performed by M.M. All authors revised and approved the final draft.

## Compliance with Ethical Standards

### Research involving Human Participants and/or Animals

This study was approved by the Institutional Review Board (IRB) of Chulalongkorn University, Bangkok, Thailand (IRB no. 62/073), which complies with the International Guideline for Human Research Protection as required by the Declaration of Helsinki.

### Informed consent

Before taking part in the study, all participants and/or their caregivers provided written informed consent.

---

**Figure 1.** Results of neural network analysis showing the relevance chart. This bargraph shows the (relative) relevance of the explanatory variables predicting stroke vs controls.

zCMPAase-zAREase: z-unit-based composite score of z chloromethyl phenylacetate (zCMPA)-ase – z arylesterase (zAREase), HT: hypertension, zCMPAase+zHDL: z-unit-based composite score of zCMPAase – z high density lipoprotein cholesterol (zHDL), BMI: body mass index

**Figure 2** Partial regression of the baseline National Institutes of Health Stroke Scale (NIHSS) score on the z-unit-based composite score of z residualized (res, after covarying for the Q192R genotype) chloromethyl phenylacetate-ase (zresCMPAase) – z res arylesterase (zresAREase) score.

**Figure 3** Partial regression of the baseline modified Rankin score (mRS) score on the z-unit-based composite score of z chloromethyl phenylacetate (CMPA)ase + z high density lipoprotein cholesterol (zHDLc), after controlling for age, sex, body mass index and Q192R genotype.

**Figure 4** Results of Partial Least Squares (PLS) path analysis. All variables were entered as single indicators except the phenome of stroke, which was constructed as a factor extracted from the baseline National Institutes of Health Stroke Scale (NIHSS) score and modified Rankin score (mRS) and an ordinal grouping as controls, and mild and moderate AIS.

This model shows that the effects of the paraoxonase (PON1) Q192R genotype on the stroke phenome are mediated by PON1 enzymatic activities, including arylesterase (AREase) and chloromethyl phenylacetate (CMPAase) activities, and a complex formed by CMPAase + high density lipoprotein cholesterol (HDLc). CRP: C-reactive protein, HT: hypertension.

Shown are the significant path coefficients (with p values) and loadings (with p values) on the extracted phenome vector. Figures within the blue circles denote the explained variance.

